# Emotional consequences of COVID-19 home confinement: The ECLB-COVID19 multicenter study

**DOI:** 10.1101/2020.05.05.20091058

**Authors:** Achraf Ammar, Patrick Mueller, Khaled Trabelsi, Hamdi Chtourou, Omar Boukhris, Liwa Masmoudi, Bassem Bouaziz, Michael Brach, Marlen Schmicker, Ellen Bentlage, Daniella How, Mona Ahmed, Asma Aloui, Omar Hammouda, Laisa Liane Paineiras-Domingos, Annemarie Braakman-jansen, Christian Wrede, Sophia Bastoni, Carlos Soares Pernambuco, Leonardo Mataruna, Morteza Taheri, Khadijeh Irandoust, Aïmen Khacharem, Nicola L Bragazzi, Karim Chamari, Jordan M Glenn, Nicholas T Bott, Faiez Gargouri, Lotfi Chaari, Hadj Batatia, Gamal Mohamed Ali, Osama Abdelkarim, Mohamed Jarraya, Kais El Abed, Nizar Souissi, Lisette Van Gemert-Pijnen, Stephen J Bailey, Wassim Moalla, Jonathan Gómez-Raja, Monique Epstein, Robbert Sanderman, Sebastian Schulz, Achim Jerg, Ramzi Al-Horani, Taysir Mansi, Mohamed Jmail, Fernando Barbosa, Fernando Santos, Boštjan Šimunič, Rado Pišot, Donald Cowan, Andrea Gaggioli, Jürgen Steinacker, Laurel Riemann, Bryan L Riemann, Notger Mueller, Tarak Driss, Anita Hoekelmann

## Abstract

**Background:** Public health recommendations and government measures during the COVID-19 pandemic have enforced restrictions on daily living, which may include social distancing, remote work/school, and home confinement. While these measures are imperative to abate the spreading of COVID-19, the impact of these restrictions on mental health and emotional wellbeing is undefined. Therefore, an international online survey was launched on April 6, 2020 in seven languages to elucidate the impact of COVID-19 restrictions on mental health and emotional well-being. This report presents the preliminary results from the first thousand responders on mental wellbeing and mood and feelings questionnaires.

**Methods:** The ECLB-COVID19 electronic survey was designed by a steering group of multidisciplinary scientists and academics, following a structured review of the literature. The survey was uploaded and shared on the Google online survey platform. Thirty-five research organizations from Europe, North-Africa, Western Asia and the Americas promoted the multi-languages survey through their networks to general society. Of the 64 questions, 7 were from the Short Warwick-Edinburgh Mental Well-being Scale (SWEMWBS), and 13 were from the Short Mood and Feelings Questionnaire (SMFQ), which are both validated instruments.

**Results:** Analysis was conducted on the first 1047 replies (54% women) from Asia (36%), Africa (40%), Europe (21%) and other (3%). The COVID-19 home confinement had a negative effect on both mental wellbeing and on mood and feelings. Specifically, a significant decrease (p<0.001 and **Δ%=** 9.4 %) in the total score of mental wellbeing was noted. More individuals (+12.89%) reported a low mental wellbeing “during” compared to “before” home confinement. Furthermore, results from the mood and feelings questionnaire (i.e., depressive symptoms) showed a significant increase by 44.9% (p<0.001) in total score with more people (+10%) developing depressive symptoms “during” compared to “before” home confinement.

**Conclusion:** The ECLB-COVID19 survey revealed an increased psychosocial strain triggered by the enforced home confinement. To mitigate this high risk of mental disorders and to foster an Active and Healthy Confinement Lifestyle (AHCL), a crisis-oriented interdisciplinary intervention is urgently needed.

## Introduction

An unexplained severe respiratory infection detected in Wuhan City of Hubei Province of China was reported to the World Health Organization (WHO) office in China on December 31, 2019. The WHO announced that the disease is caused by a new coronavirus, called COVID-19, which is the acronym of “coronavirus disease 2019”.^1^ This new virus has quickly spread worldwide. As of 14 April 2020, a total of 1.910.507 confirmed cases globally with 123.348 deaths had been reported by WHO.^2^ Considering the challenges imposed by the COVID-19 pandemic to health care systems and society in general, and in order to cut the rate of new infections and flatten the COVID-19 contagion curve, the majority of countries worldwide imposed mass home-confinement directives, with most including quarantine and social isolation.^3^ Quarantine and social isolation can be major stressors that can contribute to widespread emotional distress, as well as lead to other unexpected effects on mental health.^3–5^

Mental health is an essential component of public health because better mental health is associated with a reduced risk of several chronic diseases (e.g. dementia, depression, obesity, coronary heart disease), premature morbidity, and functional decline.^6–8^ According to the WHO, mental health is “a state of well-being in which the individual realizes his or her own abilities, can cope with the normal stresses of life, can work productively and fruitfully, and is able to make a contribution to his or her community”.^9^ There are many important facets to mental health such as personal freedoms, financial security, social stability and individual lifestyle factors (e.g. physical activity). Unfortunately, many of the social and individual consequences of the COVID-19 pandemic impose upon these facets. For example, the uncertainty of prognosis, seclusion as a result of quarantine, and financial losses associated with a reduction in economic activity likely result in several severe emotional reactions (e.g. distress) and unhealthy behaviors (e.g. excessive substance use). In this context, a recent review by Brooks et al.^10^ reported negative psychological effects, including depression, stress, fear, confusion, and anger, in quarantined people during previous epidemic. Specifically, infringement upon personal freedoms, duration of confinement, resulting financial losses, and insufficient medical care have all been suggested to increase risk for psychiatric illness during quarantine.^4^ This notion, the negative effects of quarantine on mental health including psychological and emotional problems (e.g., depression and anxiety), is directly supported by earlier studies during several outbreaks of previous infections (e.g. SARS).^11,12^

In contrast to the above earlier investigation of relatively recent infections, the dimension of the current COVID-19 pandemic drastically exceeds the previous quarantine measures, as well as the financial hardships, on an international scale. In this regard, there resides the chance of a secondary public mental health sequela related to the impact of COVID19 that extends beyond the immediate physical health crises suggesting the need to investigate the effects of COVID-19 home confinement on mental health in detail. Therefore, an international online survey (ECLB-COVID19) was launched in April 6, 2020 in multiple languages to elucidate the emotional consequences of COVID-19 home confinement. This study is the first translational large-scale survey on mental health and emotional wellbeing in the general population during the COVID-19 pandemic. It can be assumed that the COVID-19 pandemic will have essential negative implications for individual and collective mental health.

## Method

We report findings on the first 1047 replies to an international online-survey on mental health and multi-dimension lifestyle behaviors during home confinement (ECLB-COVID19). ECLB-COVID19 was opened on April 1, 2020, tested by the project’s steering group for a period of 1 week, before starting to spread it worldwide on April 6, 2020. Thirty-five research organizations from Europe, North-Africa, Western Asia and the Americas promoted dissemination and administration of the survey. ECLB-COVID19 was administered in English, German, French, Arabic, Spanish, Portuguese, and Slovenian languages. The survey included sixty-four questions on health, mental wellbeing, mood, life satisfaction and multidimension lifestyle behaviors (physical activity, diet, social participation, sleep, technology-use, need of psychosocial support). All questions were presented in a differential format, to be answered directly in sequence regarding “before” and “during” confinement conditions. The study was conducted according to the Declaration of Helsinki. The protocol and the consent form were fully approved (identification code: 62/20) by the Otto von Guericke University Ethics Committee.

### Survey development and promotion

The ECLB-COVID19 electronic survey was designed by a steering group of multidisciplinary scientists and academics (i.e., human science, sport science, neuropsychology and computer science) at the University of Magdeburg (principal investigator), the University of Sfax, the University of Münster and the University of Paris-Nanterre, following a structured review of the literature. The survey was then reviewed and edited by 50 colleagues and experts worldwide. The survey was uploaded and shared on the Google online survey platform. A link to the electronic survey was distributed worldwide by consortium colleagues via a range of methods: invitation via e-mails, shared in consortium’s faculties official pages, ResearchGate™, LinkedIn™ and other social media platforms such as Facebook™, WhatsApp™ and Twitter™. Public were also involved in the dissemination plans of our research through the promotion of the ECLB-COVID19 survey in their networks. The survey included an introductory page describing the background and the aims of the survey, the consortium, ethics information for participants and the option to choose one of seven available languages (English, German, French, Arabic, Spanish, Portuguese, and Slovenian). The present study focuses on the first thousand responses (i.e., 1047 participants), which were reached on April 11, 2020, approximately one-week after the survey began. This survey was open for all people worldwide aged 18 years or older. People with cognitive decline are excluded.

### Data privacy and consent of participation

During the informed consent process, survey participants were assured all data would be used only for research purposes. Participants’ answers are anonymous and confidential according to Google’s privacy policy (https://policies.google.com/privacy?hl=en). Participants don’t have to mention their names or contact information. In addition, participant can stop participating in the study and can leave the questionnaire at any stage before the submission process and their responses will not be saved. Response will be saved only by clicking on “submit” button. By completing the survey, participants are acknowledging the above approval form and are consenting to voluntarily participate in this anonymous study. Participants have been requested to be honest in their responses.

### Survey questionnaires

The ECLB-COVID19 is a translational electronic survey designed to assess emotional and behavioral change associated with home confinement during the COVID-19 outbreak. Therefore, a collection of validated and/or crisis-oriented brief questionnaires were included. In this manuscript, we report only results on mental wellbeing [Short Warwick-Edinburgh Mental Well-being Scale (SWEMWBS) and mood and feeling (Short Mood and Feelings Questionnaire (SMFQ)].

#### The Short Warwick-Edinburgh Mental Well-being Scale (SWEMWBS)

The SWEMWBS is a short version of the Warwick–Edinburgh Mental Wellbeing Scale (WEMWBS). The WEMWBS was developed to enable the monitoring of mental wellbeing in the general population and in response to projects, programmes and policies focusing on mental wellbeing. The SWEMWBS uses seven of the WEMWBS’s 14 statements about thoughts and feelings, which relate more to functioning than feelings suggesting an ability to detect clinically meaningful change.^13,14^ The seven statements are positively worded with five response categories from ‘none of the time (score 1)’ to ‘all of the time (score 5)’. The SWEMWBS has been recently validated for the general population and is scored by first summing the scores for each of the seven items, which are scored from 1 to 5.^15^ The total raw scores are then transformed into metric scores using the SWEMWBS conversion table. Total scores range from 7 to 35 with higher scores indicate higher positive mental wellbeing. As the idea of wellbeing is fairly new, it was suggested that further interpretation can be made depending on the study design. In this study, we considered that a score between 7 and13 reflects very low positive mental wellbeing, 14-20 reflects low positive mental wellbeing, 21-27 reflects medium positive mental wellbeing; and 28-35 reflects high positive mental wellbeing.

#### The Short Mood and Feelings Questionnaire (SMFQ)

The SMFQ is a short version of the Mood and Feelings Questionnaire (MFQ) developed by Angold and Costello in 1987.^16^ The SMFQ was developed in response to the need for a brief depression measure.^17^ The SMFQ is, therefore, suggested as a brief screening tool for depression based on thirteen of the MFQ’s 33 statements about how the subject has been feeling or acting recently.^18^ The MFQ is scored by summing together the point values of responses for each item (“not true” = 0 points; “sometimes true” = 1 point; “true” = 2 points) with higher scores on the SMFQ suggesting more severe depressive symptoms. Scores on SMFQ range from 0 to 26. A total score of 12 or higher may indicate the presence of depression in the respondent.^18^

### Data Analysis

Descriptive statistics were used to define the proportion of responses for each question and the distribution of the total score of both questionnaires. All statistical analyses were performed using the commercial statistical software STATISTICA (StatSoft, Paris, France, version 10.0) and Microsoft Excel 2010. Normality of the data distribution in each question was confirmed using the Shapiro-Wilks-W-test. Values were computed and reported as mean ± SD (standard deviation). To assess for significant differences in responses “before” and “during” the confinement period, paired samples t-tests were used for normally distributed data (responses to the *SWEMWBS* questionnaire) and the Wilcoxon test was used when normality was not assumed (responses to the SMFQ). Effect size (Cohen’s d) was calculated to determine the magnitude of the change of the score and was interpreted using the following criteria: 0.2 (small), 0.5 (moderate), and 0.8 (large)^19^ Statistical significance was accepted as α<0.05.

## Results

### Sample description

The present study focused on the first thousand responses (i.e., 1047 participants), which was reached on April, 11, 2020. Overall, 54% of the participants were women, and the participants were from Western Asia (36%), North Africa (40%), Europe (21%) and other (3%). Age, health status, employment status, level of education and marital status are presented in Table 1.

**Table 1.** Demographic characteristics of the participants.

| Variables | N | (%) |
| --- | --- | --- |
| **Gender** |  |  |
| Male | 484 | (46.2%) |
| Female | 563 | (53.8%) |
| **Continent** |  |  |
| North Africa | 419 | (40%) |
| Western Asia | 377 | (36%) |
| Europe | 220 | (21%) |
| Other | 31 | (3%) |
| **Age (years)** |  |  |
| 18-35 | 577 | (55.1%) |
| 36-55 | 367 | (35.1%) |
| >55 | 103 | (9.8%) |
| **Level of Education** |  |  |
| Master/doctorate degree | 527 | (50.3%) |
| Bachelor's degree | 397 | (37.9%) |
| Professional degree | 28 | (2.7%) |
| High school graduate, diploma or the equivalent | 69 | (6.6%) |
| No schooling completed | 26 | (2.5%) |
| **Marital status** |  |  |
| Single | 455 | (43.4%) |
| Married/Living as couple | 562 | (53.7%) |
| Widowed/Divorced/Separated | 30 | (2.9%) |
| **Employment status** |  |  |
| Employed for wages | 538 | (51.4%) |
| Self-employed | 74 | (7.1%) |
| Out of work/Unemployed | 75 | (7.2%) |
| A student | 259 | (24.7%) |
| Retired | 23 | (2.2%) |
| Unable to work | 9 | (0.85%) |
| Problem caused by COVID-19 | 59 | (5.6%) |
| Other | 10 | (0.95%) |
| **Health state** |  |  |
| Healthy | 956 | (91.3%) |
| With risk factors for cardiovascular disease | 81 | (7.7%) |
| With cardiovascular disease | 10 | (1%) |

### The Short Warwick-Edinburgh Mental Well-being Scale (SWEMWBS)

Change in mental well-being score assessed through the SWEMWBS from “before” to “during” confinement period are presented in Table 2. The total score decreased significantly by 9.4% during compared to before home confinement (t=18.82, p<0.001, d=0.58). A statistically significant decrease was observed for each of the 7 questions. Particularly, feeling related questions such as feeling optimistic, useful, relaxed and close to others showed a lower score at “during” compared to “before” confinement with |Δ%| ranged from 4% to 13% (3.44 ≤ t ≤ 20.26; P < 0.001; 0.106 ≤ d ≤ 0.626). Similarly, participants scored lower in thinking related questions “during” compared to “before” confinement period with |Δ%| ranged from 7% to 16% for the capacities to deal well with problems, think clearly and make up own mind about things (10.36 ≤ t ≤ 12.89 ≤ P < 0.001, 0.32 ≤ d ≤ 0.51).

**Table 2.** Responses to the Short Warwick-Edinburgh Mental Well-being Scale before and during home confinement.

| Questions | Before<br>confinement | During<br>confinement | $\Delta$ ($\Delta\%$) | 95% IC | t test | p<br>value | Cohen's<br>d |
| --- | --- | --- | --- | --- | --- | --- | --- |
| 1. I've been feeling optimistic about the future | 4.08±0.91 | 3.54±1.11 | -0.54 (-13.2%) | 0.49 - 0.59 | 20.260 | <.001 | 0.626 |
| 2. I've been feeling useful | 4.05±0.89 | 3.62±1.13 | -0.43 (-10.7%) | 0.37 - 0.49 | 14.605 | <.001 | 0.451 |
| 3. I've been feeling relaxed | 3.38±0.94 | 3.25±1.07 | -0.13 (-3.9%) | 0.06 - 0.21 | 3.442 | <.001 | 0.106 |
| 4. I've been dealing with problems well | 3.88±0.81 | 3.62±0.93 | -0.26 (-6.6%) | 0.21 - 0.3 | 10.749 | <.001 | 0.332 |
| 5. I've been thinking clearly | 3.99±0.77 | 3.71±0.94 | -0.28 (-6.9%) | 0.22 - 0.33 | 10.368 | <.001 | 0.320 |
| 6. I've been feeling close to other people | 3.88±0.92 | 3.26±1.16 | -0.61 (-15.8%) | 0.54 - 0.69 | 16.644 | <.001 | 0.514 |
| 7. I've been able to make up my own mind about things | 4.04±0.83 | 3.72±1.00 | -0.32 (-7.9%) | 0.27 - 0.37 | 12.887 | <.001 | 0.398 |
| Total score | 27.3±4.37 | 24.73±5.18 | -2.57 (-9.4%) | 2.3 - 2.84 | 18.821 | <.001 | 0.582 |

### The Short Mood and Feelings Questionnaire (SMFQ)

Change in mood and feeling score from “before” to “during” confinement period in response to SMFQ depression monitoring tool are presented in Table 3. The SMFQ total score increased significantly by 44.9% “during” compared to “before” home confinement (z=14.52, p<0.001, d=0.44). For most questions, an increased score was noted with the following exceptions: “I was a bad person” and “I did everything wrong”. Particularly, bad-feeling related questions such as unhappy, unenjoyed, tired, hated himself, no good and lonely, showed higher score at “during” compared to “before” confinement with |Δ%| ranged from 37% to 107% (5.07 ≤ z ≤ 12.60; P < 0.001, 0.17 ≤ d ≤ 0.47). Similarly, scored responses to questions related to how the subject has been acting (i.e., restless, crying and doing nothing) or thinking (i.e., not properly, not concentrated, unloved and not good as others) in bad way showed higher score at “during” compared to “before” confinement with |Δ%| ranged from 10% to 76% (2.30 ≤ z ≤ 9.82; 0.45 ≤ P ≤ 0.001, 0.07 ≤ d ≤ 0.46).

**Table 3.** Responses to the Short Mood and Feelings Questionnaire before and during home confinement.

| Questions | Before<br>confinement | During<br>confinement | $\Delta$ ($\Delta\%$) | z values | 95% IC | p<br>value | Cohen's<br>d |
| --- | --- | --- | --- | --- | --- | --- | --- |
| 1. I felt miserable or unhappy | 0.49±0.57 | 0.79±0.72 | 0.30 (61.2%) | z=12.124 | -0.34 - 0.26 | <.001 | 0.458 |
| 2. I didn't enjoy anything at all | 0.29±0.51 | 0.6±0.7 | 0.31 (107.7%) | z=12.609 | -0.35 - 0.27 | <.001 | 0.468 |
| 3. I felt so tired I just sat around and did nothing | 0.46±0.6 | 0.81±0.78 | 0.35 (76.2%) | z=12.456 | -0.39 - 0.3 | <.001 | 0.460 |
| 4. I was very restless | 0.46±0.6 | 0.66±0.75 | 0.20 (44%) | z=7.762 | -0.25 - 0.16 | <.001 | 0.271 |
| 5. I felt I was no good anymore | 0.34±0.53 | 0.55±0.71 | 0.21 (62.3%) | z=9.822 | -0.25 - 0.18 | <.001 | 0.351 |
| 6. I cried a lot | 0.39±0.6 | 0.43±0.67 | 0.04 (10.1%) | z=1.997 | -0.07 - 0.01 | 0.045 | 0.071 |
| 7. I found it hard to think properly or concentrate | 0.53±0.58 | 0.77±0.74 | 0.24 (45.1%) | z=9.370 | -0.28 - 0.20 | <.001 | 0.336 |
| 8. I hated myself | 0.23±0.49 | 0.32±0.6 | 0.09 (37.3%) | z=5.074 | -0.12 - 0.06 | <.001 | 0.175 |
| 9. I was a bad person | 0.15±0.39 | 0.17±0.44 | 0.01 (8.6%) | z=1.121 | -0.04 - 0.01 | 0.262 | 0.037 |
| 10. I felt lonely | 0.39±0.58 | 0.59±0.73 | 0.2 (52.2%) | z=8.740 | -0.24 - -0.16 | <.001 | 0.308 |
| 11. I thought nobody really loved me | 0.26±0.52 | 0.29±0.57 | 0.03 (10.2%) | z=2.296 | -0.05 - 0.01 | 0.021 | 0.080 |
| 12. I thought I could never be as good as other people | 0.23±0.49 | 0.26±0.54 | 0.04 (16.4%) | z=3.152 | -0.06 - 0.02 | <.001 | 0.108 |
| 13. I did everything wrong | 0.27±0.49 | 0.27±0.49 | 0.0 (0.3%) | z=0.080 | -0.02 - 0.02 | 0.936 | 0.002 |
| Total score | 4.49±4.41 | 6.5±5.63 | 2.01 (44.9%) | z=14.520 | -2.29 - -1.73 | <.001 | 0.436 |

## Discussion

The present study reports results from the first 1047 participants (54% female) who responded to our ECLB-COVID19 multiple languages online survey. Findings indicate significant negative effects of the current Covid-19 pandemic on mental health, especially mental wellbeing, mood, and feeling. There, mental wellbeing (estimate with the total score in SWEMWBS) decreased significantly by 9.4 % during home confinement with more individuals (+12.89%) reporting a very low to low mental wellbeing at “during” compared to “before” home confinement. The largest effects of the current COVID-19 pandemic were observed in questions related to optimistic feeling, closed to others, useful, and thinking. Furthermore, results from the mood and feelings questionnaire showed significant increase by 44.9% in SMFQ total score, indicating negative effects with more people (+10%) developing depressive symptoms at “during” compared to “before” home confinement. Especially, questions related to unhappiness, unenjoyment, bad feeling, unclear thinking and loneliness showed highest effect sizes.

The present findings support previous reports suggesting several psychological perturbations and mood disturbances such as stress, depression, irritability, insomnia, fear, confusion, anger, frustration, boredom, and stigma during quarantine periods of earlier infection.^10,20,21^ Regarding the COVID-19 related research, first results from Chinese studies indicate that the COVID-19 outbreak engendered anxiety, depression, sleep problems, and other psychological problems.^22,23^ The significantly lower total SWEMWBS score and higher total SMFQ score “during” compared to “before” confinement support the negative effects of the current COVID-19 pandemic on mental wellbeing and emotional state in participants from Western Asian, North Africa and Europe. Taken together, findings from China and from our survey provide insight into the risk of worldwide emotional distress and mental functioning (e.g. low wellbeing, anxiety, depression) during the COVID-19 home confinement period.

Weakening of physical and social contacts with the disruption of normal lifestyles (lower freedoms, financial losses, sedentariness, sleep disorder, unhealthy diet, etc.) during the COVID-19 outbreaks, have been suggested as major risk factors for lower emotional wellbeing and mental disorders.^5,24^ Furthermore, research indicates that some groups may be more vulnerable to the psychosocial effects of the COVID-19 pandemic. Particularly, people with risk factors for COVID-19 infection (e.g. diabetes, chronic heart failure, COPD, immune deficiency), people living in congregate settings (e.g. Hospice) and people with a predisposition and/or pre-existing psychiatric or substance use problems are at increased risk for mental health problems.^4^

Since mental disorders have been previously identified as risk factors for several chronic diseases (e.g. hypertension; obesity, dementia)^8,25–27^ and showed to be associated with increased mortality,^28,29^ a crisis-oriented interdisciplinary intervention approach to promote wellbeing and mitigate the negative effects of the COVID-19 pandemic on mental health is urgently needed.^30,32^ Given that an active lifestyle including physical and social activity is an important modifiable factor for mental health across the lifespan,^33^ this intervention should focus on fostering social communication and physical activity. In this context, it is recommended that stakeholders encourage individuals (i) to promote their social participation through technology (e.g., social platform, gamification, video chat, group conversation etc.), and (ii) to engage in indoor and/or outdoor physical activity in large public parks, whilst conforming with distancing and hygiene recommendations. Furthermore, for the more vulnerable population to the psychosocial strain, supportive intervention should also include “need-oriented” psychosocial services (e.g., psychoeducation, cognitive behavioural techniques, and/or consulting with specialists) delivered by means of telemedicine.

However, to ensure a sustainable intervention approach, future research should investigate the long-term impact of the COVID-19 pandemic on mental health and identify which component(s) of psychosocial strain may persist after the quarantine.

## Strength and limitation

The strength of this study is that the data was collected very quickly during the restrictions using a fully anonymous cross-disciplinary survey provided in multiple language and widely distributed in several continents. However, given that most participants (90.2%) were 55 years old or younger, were healthy (90.5%), and were educated (90.9%) with a degree beyond high school. These demographic characteristics may relate to the results obtained and, thus, the present findings need to be interpreted with caution. Additionally, since the ECLB-COVID19 survey is still open and meanwhile also available in Dutch, Persian and Italian languages, future post-hoc studies in a more representative sample will be conducted to assess the interaction between the mental and emotional strain evoked by COVID-19 and the demographical characteristics of the participants.

## Conclusion

Besides stresses inherent in the illness itself, results from the ECLB-COVID19 survey reveal a negative effect of home-confinement on mental and emotional wellbeing with more people developing depressive symptoms “during” compared to “before” the confinement period. This increased psychosocial strain triggered by the enforced home confinement should encourage stakeholders and policy makers to implement a crisis-oriented interdisciplinary intervention to mitigate the negative effects of restrictions and to foster an Active and Healthy Confinement Lifestyle (AHCL).

## Data Availability

Data are available by the corresponding author upon reasonable request

## Acknowledgments

We thank our consortium’s colleagues who provided insight and expertise that greatly assisted the research. We thank all colleagues and peoples who believed on this initiative and helped to distribute the anonymous survey worldwide. We are also immensely grateful to all participants who #StayHome & #BoostResearch by voluntarily taken the #ECLB-COVID19 survey.

## Funding

Research are urgently needed to help understand the impacts of the covid-19 pandemic on peoples’ lifestyle. However, normal funding mechanism to support scientific research are too slow. The authors received no specific funding for this work.

## Conflicts of Interest

The authors report no conflicts of interest, no relevant disclosures and no external financial support. The authors alone are responsible for the content and writing of the paper.

## Notes

### Competing Interest Statement

The authors have declared no competing interest.

